# ‘On Your Mark’: Operationalizing a Readiness for Change Module in the SHIFT Intervention

**DOI:** 10.1101/2025.10.09.25337684

**Authors:** Liane R. Ginsburg, Whitney Berta, Matthias Hoben, Frode F. Jacobsen, Lonnie Kehler, Jennifer Pietracci, Laurel Rose, Danielle Saj, Georgina Veldhorst, Adrian Wagg, Malcolm Doupe

**Author notes:** **Address correspondence to:** Liane Ginsburg, Professor, School of Health Policy & Management, York University, Stong College 353, 4700 Keele Street, Toronto, ON M3J 1P3 Canada.

## Abstract

**Background:** Readiness for change (R4C) is an important antecedent of care practice change and is linked to a range of implementation and effectiveness outcomes. We describe the creation, piloting, and evaluation of R4C activities intended to help nursing home (NH) teams implement a quality improvement intervention.

**Methods:** We developed and operationalized a R4C module with activities conducted with NH leaders during intervention recruitment. An implementation pilot was conducted in three NHs and evaluated using a mixed methods process evaluation guided by Proctor’s (2011) implementation outcomes framework.

**Results:** R4C activities were feasible, acceptable, and delivered with fidelity. The approach promoted readiness among senior leaders, but not among front-line teams. Results also showed (1) R4C activities are closely tied to other variables (e.g., NH leadership facilitation) that influence implementation, and (2) core R4C components (i.e., change efficacy) can emerge during an intervention, pending teams’ perceived success

**Discussion:** We suggest conducting R4C activities with all of those involved *across an organization* who have a role to play in change implementation. Findings also reveal challenges associated with isolating the effects of ‘pre-intervention’ R4C activities on intervention implementation and success. Pilot results will inform a larger-scale quality improvement study.

**Trial registration:** Registered at ClinicalTrials.gov (ID NCT03426072) on July 18, 2022.

**KEY MESSAGES REGARDING FEASIBILITY:**

1. What uncertainties existed regarding the feasibility?
  - While ‘readiness for change’ has been identified as an important lever for implementation of evidence-informed practice change, there is little in the published literature regarding how to operationalize of R4C to promote uptake of practice change initiatives. There is also uncertainty regarding ‘implementability’ (feasibility, acceptability, appropriateness, fidelity) of R4C activities in complex care settings
2. What are the key feasibility findings?
  - Up-front activities requiring a few hours of a facility leader’s time, designed to gauge and also operationalize and promote readiness for change were both feasible and acceptable and were delivered with fidelity. Facility leaders showed less fidelity *enacting* the intervention support and communication roles targeted by the R4C activities. A ‘broken telephone’ scenario existed where senior leaders did not convey to unit leaders what their project roles entailed.
3. What are the implications of the feasibility findings for the design of the main study?
  - Our results suggest that it is necessary to actively target R4C activities directly to all organizational members involved in the practice change initiative. Failure to promote linkages and role clarity across participants at different levels of an organization makes it likely that change actors will remain unsure of their role in the change initiative. The design of the main study ultimately included two additional R4C focused meetings (one to bring senior and team sponsors together, one to bring team sponsors and team members together) to promote greater role clarity and enhanced project preparation.

## BACKGROUND

While traditional experimental designs primarily seek to conclude whether interventions/innovations are effective, the goal of implementation science goes beyond that. Engaging with decision makers [1,2], implementation science aims to make interventions work in complex settings – ultimately to promote their sustainment (of actions), sustainability (of outcomes), and eventual scale up. Concerned with questions such as “how to successfully implement interventions into practice”, implementation science research has largely explored key *determinants* of implementation [3]. To promote greater knowledge uptake, attention to more dynamic questions regarding implementation *processes*, and their key determinants, is required [4–6].

Among the determinants of greater research uptake, the literature suggests *readiness for change* is important. Readiness for change (R4C) is a “state of preparedness for future action”[7] – a state that is both psychological and structural [8]. R4C is a “multi-level construct” that should be considered at the individual, group, and organizational levels [8,9]. It reflects both the willingness (commitment) and ability (efficacy) to support and implement change [8]. R4C has been shown to impact the implementation of a range of practice changes [10,11]. By operationalizing R4C and exploring its role in the uptake of one such initiative, ‘Supporting Healthcare Improvement Through Facilitation and Training’ (SHIFT), this paper contributes to a small but growing body of literature on how to promote/build readiness for change [12,13] at the start of a practice change initiative.

### The Program – From SCOPE to SHIFT

Quality improvement (QI) interventions in healthcare aim to improve patient or staff outcomes, achieve more effective use of resources, and/or improve processes of care [14]. The ultimate goal of QI interventions is to introduce large-scale changes (i.e., spread across multiple patients, staff, sites) that are sustained once the research-intensive phase of the intervention is complete [1].

The ‘**S**afer **C**are for **O**lder **P**ersons (in residential) **E**nvironments’ (SCOPE) intervention was based on a compendium of implementation science activities conducted as part of the long-standing Translating Research in Elder Care (TREC) research program conducted in nursing homes across Western Canada [15,16]. SCOPE knowledge has been reported through a proof of principle study [17], a pilot [18], a full trial [19], and process evaluation work [5,6,20]. SCOPE utilized a modified version of the Institute for Healthcare Improvement (IHI) Breakthrough Series Collaborative [21]. SCOPE uses facilitated support to empower healthcare aide (HCA) teams to conduct QI initiatives to improve their work life environment (e.g., by fostering empowerment and building more effective care teams), and ultimately improve the quality of care for residents [17–19]. The core components of the SCOPE intervention include (i) use of the IHI Plan-Do-Study-Act (PDSA) Model for Improvement as the core “engine for change”, (ii) quarterly in-person learning congresses (LCs) attended by all participating teams, (iii) ongoing support from an external Quality Advisor (QA) during facilitated action periods between LCs, and (iv) additional QA training provided to unit and facility leaders to help them support frontline teams. Importantly, SCOPE frontline teams were led by, and comprised mostly of, HCAs given the major role they play in providing direct care to NH residents in Canada [22] and their unique capacity to understand and respond to residents’ daily needs.

SCOPE results showed that teams were committed to improving quality; experienced pride, empowerment, feelings of change efficacy, and believed that the intervention led to improvements in resident care; and teams felt that they forged more trusting relationships with staff in other disciplines and with managers [5,6,20]. The challenges encountered in SCOPE involved navigating roles, achieving buy-in from non-SCOPE colleagues, carrying out measurement aspects of the PDSA improvement cycle, and finding time to plan and carry out improvement plans [5,6].

To maintain SCOPE successes and address these challenges, SHIFT augments the core SCOPE components by (i) more purposefully preparing teams through a readiness for change (R4C) module, and (ii) designing and implementing a leadership development module (LeaderSHIFT) to help unit-level leaders more effectively support front-line teams implementing the QI intervention.

### Objectives

While R4C is recognized as important to the implementation process, we lack detailed knowledge regarding both the strategies to operationalize key R4C principles, and the consequences of integrating these principles into intervention plans (e.g., do R4C activities enable teams to implement their quality improvement ideas sooner or more completely?). This manuscript has two feasibility-related[36] objectives:

1. to describe a module of activities we developed to operationalize (i.e., gauge and promote) R4C;
2. to present results from a pilot implementation study which enable us to refine these R4C activities for subsequent application in the full SHIFT intervention.

To help orient the reader, Figure 1 shows the structure of the final (i.e., post-pilot) SHIFT intervention, with R4C activities (module 1) highlighted in the top section. The full SHIFT program shown in Figure 1 and the Leadership module (LeaderSHIFT – module 2) are described in two other manuscripts currently under review.

**Figure 1.**
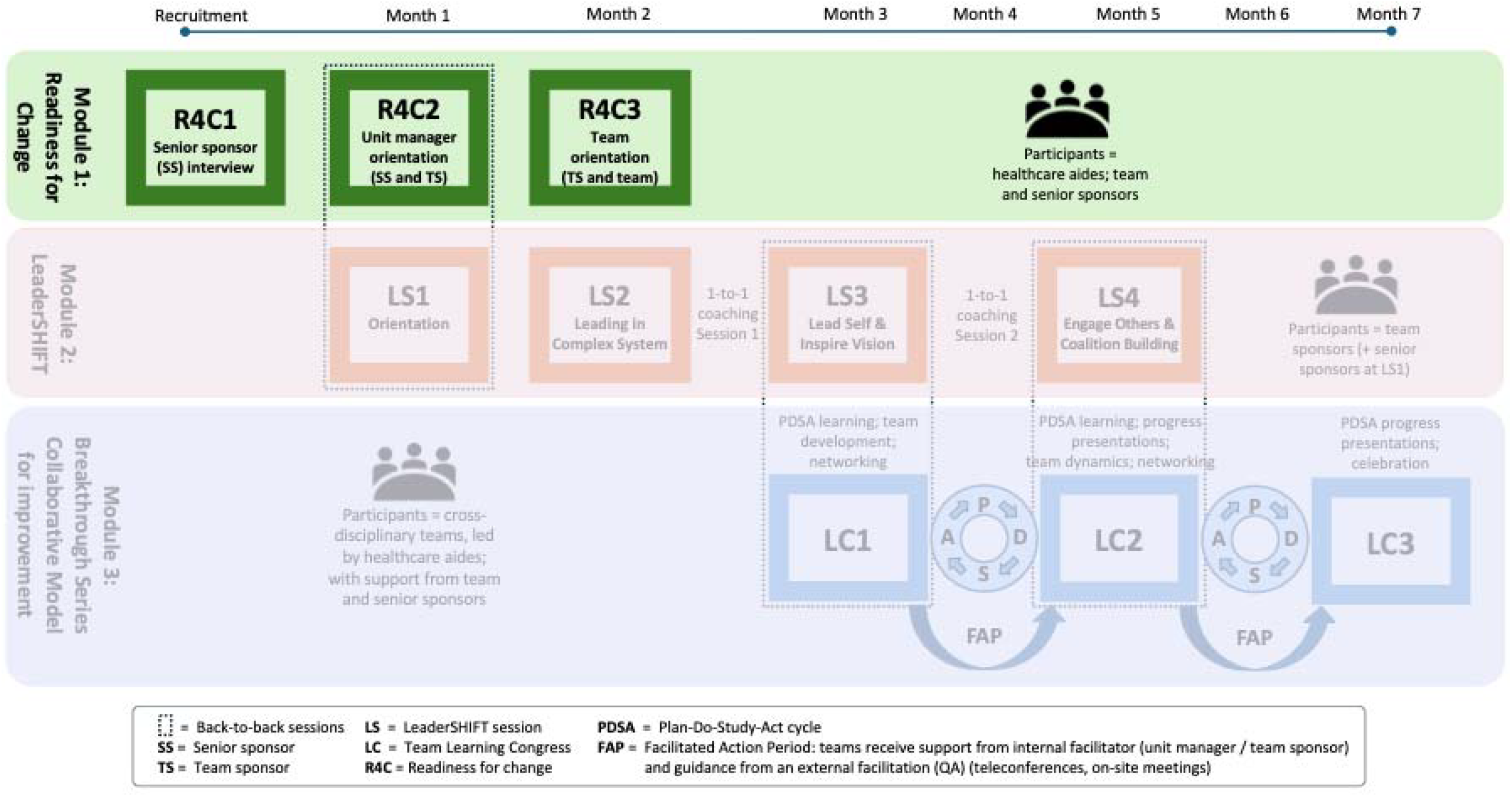
SHIFT Program Structure.

## METHODS

Initial R4C activities were guided by (i) theoretical and empirical readiness for change literature, (ii) knowledge from previous SCOPE initiatives described above, and (iii) collaborations with local health system partners reflecting implementation science principles of stakeholder codesign and feasibility (e.g., to ensure tasks were feasible, had value, and were not overtly onerous from the perspective of our partners). The R4C approach was pilot tested with three nursing homes in the Canadian province of Manitoba in 2023, as part of the SHIFT pilot, and was evaluated using a mixed methods concurrent process evaluation. This study adheres to relevant sections of the Template for Intervention Description and Replication (TIDierR)[39] and CONSORT (Consolidated Standards of Reporting Trials) extension for pilot and feasibility trials[40].

### Developing the Readiness for Change Module – R4C Theories that Align with SHIFT

Organizational readiness for change will depend on the particular context of a change initiative [8]. Readiness for change elements at the organizational and individual levels relevant to SHIFT and aligned with Weiner’s conceptualization of R4C (which includes change commitment and change efficacy [8]) were identified using the Organizational Readiness to Change Assessment (ORCA)[23], and Individual Readiness for Organizational Change (IROC) concepts [24]. The ORCA was used given its theoretical roots in the PARIHS framework for research implementation [25–27] which also guided SCOPE and SHIFT and emphasizes facilitation as a key factor influencing implementation [23]. The IROC was used given its emphasis on the importance of organizational member’s “cognitive and emotional inclination to accept, embrace, and adopt” [24:235] a particular change initiative which is consistent with prior work in SCOPE [20].

Through iterative assessment of the ORCA and IROC tools, and prior empirical work on SCOPE implementation challenges described above (e.g., navigating roles, finding time to plan and carry out SCOPE improvement activities), R4C activities were developed that focused on identified contextual deficits, specifically: enriching facilitation (defined as an organization’s capacity to provide internal facilitation to support change [23]), along with strategies to improve management support, change valence, change efficacy, and change commitment. In the context of readiness for change, management support refers to the extent to which organizational members feel that senior leaders support and are committed to the proposed change [24], change valence reflects one’s belief in the value of the intended change, and may be driven by a variety of factors[8]. Change efficacy reflects one’s belief in their ability to carry out a course of action - and is important because low efficacy reduces motivation to engage in that course of action [25]. Change commitment reflects one’s willingness (i.e., their psychological / motivational readiness) to engage in a change initiative [26].

Table 1 lists the relevant ORCA and IROC elements that guided our efforts to define a R4C module and operationalize it in the SHIFT Pilot study. As noted, these R4C elements align with Weiner’s conceptualization of R4C which includes change commitment and change efficacy [8].

**Table 1.** Readiness for Change Elements Relevant to SHIFT.

| **Element** | **Source** |
| --- | --- |
| Organizational leaders and unit champion who agree with intervention goals/are committed and set the intervention as a priority | ORCA (facilitation) / IROC (management support) / Change commitment (Weiner, 2008, 2009) |
| Organizational leaders and unit champion who agree to support and resource the intervention; Team members are given time, regular meetings, and resources to accomplish intervention tasks | ORCA (facilitation) / IROC (management support) |
| Senior leaders emphasize the importance of the intervention and encourage others to embrace it | IROC (management support) / Change commitment (Weiner, 2009) |
| Unit champion who works well with others across the hierarchy | ORCA (facilitation) |
| Implementation plan outlines clear roles and responsibilities | ORCA (facilitation) |
| Team members (including leaders) are prepared to share responsibility and acknowledge input across the hierarchy | ORCA (facilitation) |
| Belief in one's ability to adjust to and manage new tasks that are part of the intervention | IROC (change efficacy) |
| Belief one has / can learn the required skills to carry out intervention tasks | IROC (change efficacy) |
| Past experiences that provide confidence to do the work | IROC (change efficacy) |
| Belief the intervention fits with organizational priorities / is justified / will benefit the organization | IROC (organizational change valence) |
| Belief the intervention will be personally valuable to individual team members | IROC (Personal change valence) |

### Promoting / Operationalizing Readiness for Change in the SHIFT Pilot

To promote readiness for change, R4C elements relevant to SHIFT (table 1) were operationalized during the initial phases of the SHIFT pilot during site recruitment, unit identification, and team orientation.

#### Site recruitment

To promote change commitment, willingness to participate was deemed to be a key factor. Sites were therefore recruited purposefully. The research team, in conversation with the regional government authority that funds and oversees healthcare (the Winnipeg Regional Health Authority) and NH site leaders, collaborated to identify two to three sites felt to have the capacity to participate. Leaders of potential sites were invited to meet virtually for a 90-minute interview with the project coordinator to both gauge and promote the R4C elements in Table 1. To promote change valence, change efficacy, senior management support and change commitment, the interview included detailed discussions regarding the benefits of project participation, project roles and responsibilities, and commitments required, respectively. Consent to participate was obtained, and a memorandum of understanding outlining roles and responsibilities of the sites and the research team was signed. The second part of the interview focused on strategies the facility leader could use to identify the most appropriate care unit/unit leader within the facility for SHIFT participation.

#### Unit identification

To promote management support and facilitation, we encouraged facility leaders to nominate themselves as a ‘senior sponsors’ and to seek unit leaders (‘team sponsors’) with the requisite leadership, interpersonal skills, and social capital to be effective in their roles. Unit managers (typically Registered Nurses, RNs) fulfilled the role of ‘team sponsor’ in SHIFT and were charged with removing obstacles for the team and providing support for the HCAs leading the team. The team sponsor is akin to a QI project champion – a role found to be important for implementation success [23]. Based on the R4C and QI champion literature [23– 25], facilities were encouraged to identify and invite unit leaders with longer tenure, who they felt had an interest in QI and openness to new ideas and care innovation, and sufficient influence and social capital to engage team members. Unit leaders were also identified based upon facility leader’s perception of their: abilities to build trust and advocate for/prioritize the intervention, openness to a distributed leadership approach, and capacity – in terms of both time and motivation – to engage with front-line staff during SHIFT. To promote R4C among team members, team sponsors were encouraged to use the same characteristics just described to guide recruitment of HCAs.

#### Team orientation

To foster change efficacy among team members, roles and responsibilities of team members, their sponsors, and the quality advisor were made clear during an initial in-person team orientation meeting facilitated by the Quality Advisor (QA). This meeting also engaged team members in selecting the clinical focus of their improvement work. Focusing teams on care-sensitive topics within HCAs’ scope of practice (topics amenable to improvement while requiring no or only minimal additional resources), and ensuring the topic was meaningful to the HCAs were strategies intended to promote change efficacy, valence, and commitment among SHIFT team members.

### Evaluating Readiness for Change in the SHIFT Pilot

While we were interested in ascertaining the extent to which the *level of readiness for change* (the outcome) was influenced by our R4C activities, we were equally interested in evaluating the *approaches we used to gauge and enhance readiness for change* within the pilot sites.

A mixed-methods concurrent process evaluation was conducted using previously employed approaches [5] including: qualitative analysis of Quality Advisor diaries, participant focus groups, and interview and observer field notes, and quantitative analysis of participant survey data (described in Table 2 below).

**Table 2.** R4C Pilot Data Collection Approaches.

|  | **Approach** | **Purpose & Evaluation Aim** | **Data Quality Approach** |
| --- | --- | --- | --- |
| Field Notes | Research staff used a semi-structured template to observe all interactions with the SHIFT teams. | To capture qualitative data on fidelity (delivery and enactment) of the R4C approach, the extent to which teams engage in the R4C discussions<sup>†</sup>, and their level of readiness<sup>††</sup> | QAs were trained using diary entry examples. Quality and completeness of initial field notes and diary entries were assessed by a researcher to ensure relevant information was captured in sufficient detail. |
| Quality Advisor Diary | QAs made a diary entry after each interaction with a team (team orientation meeting; at and between SHIFT LCs) | To capture qualitative data on acceptability, feasibility, and fidelity of the R4C approach<sup>†</sup>; as well as their level of readiness<sup>††</sup> |  |
| Initial R4C Interviews | Interview with facility senior leader to gauge and promote R4C at the start of the intervention (during site recruitment). | To obtain data directly from the facility's senior leader on their / the organization's level of R4C<sup>††</sup> | Interview was facilitated by a member of the research team familiar with qualitative methods, recorded, and transcribed. |
| Focus Groups | Conducted at the end of SHIFT with HCAs from each team to obtain *team-specific* data. A separate focus group was conducted across teams with Team Sponsors. | To obtain retrospective data directly from SHIFT participants on their / the organization's level of R4C<sup>††</sup> and the acceptability and feasibility of the R4C approach<sup>†</sup> | Adhere to principle of homogeneous strangers to eliminate power differentials within a focus group [30]. Focus groups were recorded, and transcribed. |
| Questionnaires | IROC questionnaire completed at the initial SHIFT LCs by team and senior sponsors and team members. | To collect data from multiple stakeholders regarding the organization's level of R4C<sup>††</sup> | Collected at LCs to reduce data collection burden and ensure high response rates; IROC is a validated instrument [29]. |

<sup>†</sup> to evaluate *processes for gauging and enhancing SHIFT readiness*
<sup>††</sup> to evaluate *level of readiness*.

### Evaluation of processes to gauge and enhance SHIFT readiness

Evaluation of the *processes* we used to gauge baseline SHIFT readiness and enhance it was guided by Proctor’s implementation outcomes framework [26]. We examined the framework’s three most relevant implementation outcomes: *acceptability* and *feasibility* of the R4C approach described above, and *fidelity* to it. Acceptability reflects stakeholder perception that the approach is agreeable/satisfactory while feasibility refers to the extent to which the approach can be carried out in the facility. These were important to assess, as senior leaders’ receptivity to R4C activities (e.g., balancing value with the time taken to complete) is both critical for implementation and eventual scale up. Fidelity refers to the extent to which the readiness for change approach was implemented as intended. Consistent with the Behavioural Change Consortium’s conceptualization of fidelity [27], our assessment explored *delivery* of the R4C approach to the participants as planned, and senior and team sponsor *enactment* of their roles (e.g., identifying suitable team sponsors and team members and providing program support). Acceptability, feasibility, and fidelity were assessed using observer field notes, diaries, survey questions, and an end-of-pilot focus group (see Table 2 below).

### Evaluation of Level of Readiness

To evaluate level of readiness of the three SHIFT pilot sites, we used both qualitative and quantitative approaches. R4C was assessed **qualitatively** at the start of the recruitment processes described above. The initial recruitment interview asked the senior sponsor questions to gauge (1) R4C organizational valence (e.g., “Do you think an intervention like this could help improve care for residents at your facility?”), (2) R4C change efficacy and management support (e.g., “To what extent would you be willing / able to support and be involved in the SHIFT intervention?”), and (3) facilitation aspects of R4C (e.g., “Do you have resources available to support the team members? E.g., dedicated time / staffing levels for team members to plan and work on improvement projects.”).

During the initial recruitment interview, the senior sponsor was asked questions to: (a) prompt identification of a team sponsor likely to provide management support and facilitation (e.g., “Is there a leader on one of your units who generally thinks HCAs are capable of making impactful change versus should just listen to nurses and follow tasks?”); (b) assess change efficacy (e.g, “Can you think of a leader who has the self-confidence to support the intervention)?”; and (c) assess change valence (“Can you think of a leader who believes in the importance of QI interventions”?).

Readiness for change was also assessed **quantitatively**. Because readiness is a multi-level construct and implementation requires actions organized across multiple levels of an organization [8,28], the IROC survey [29] was administered to the senior sponsor, team sponsor, and front-line team members. The instrument is used to measure readiness for organizational change at the individual level and has 25 items in 4 domains: (i) change efficacy; 6 items, (ii) change valence; 10 items, (iii) senior management support; 6 items, and (iv) personal valence; 3 items. All items use a 5-point Likert type response scale where a higher score reflects greater R4C. The IROC survey was administered at the first SHIFT LC shortly after teams were formed when all sponsors and team members were present.

Table 2 summarizes all data collection approaches used in the R4C pilot. Data were content analyzed by two of the authors (authors 1 & 5) to assess (1) R4C acceptability, feasibility and fidelity, (2) teams’ actual level of R4C, and (3) to suggest modifications to the R4C approach following the pilot.

## RESULTS

### Processes for Gauging and Enhancing SHIFT Readiness for Change

#### Acceptability and Feasibility

Analyses of the senior sponsor interviews suggest the R4C recruitment interview approach used to gauge and enhance R4C potential was both acceptable to senior sponsors and considered feasible to implement. Field notes and interview data showed that senior sponsors participated in the 90-minute R4C interview, engaged in the discussion, and ended by identifying a unit and manager that they felt would best be able to fulfil the team sponsor role in SHIFT.

#### Fidelity

The initial R4C interview was *delivered* as intended and the process enabled the senior sponsor to *enact* aspects of R4C related to selecting an appropriate unit / team sponsor. For example, in their interview, one senior sponsor said that they selected the team sponsor because,

> “They [staff] speak very openly and candidly about how much support they get from their manager…how well the communication is on the team [because of her] leadership on the unit and they see her as someone who they can trust…I see a willingness and an eagerness among the team on [unit X] that I don’t see in the same or similar measure on [unit Y].” [SS interview]

However, the end-of-pilot team sponsor focus group data indicated that team sponsors did not *enact* the process they were asked to use to recruit a HCA team to promote R4C. The information provided during the senior sponsor interview encouraged sites to choose HCAs with leadership potential and to *invite* (rather than *tell*) them to participate. When asked if they received advice on choosing team members, one team sponsor said, “I just based mine [selection] on the –being fulltime because there’s more contact with the residents, so”. Another stated: “if you pick the one that has the most influence, you know the others will – you know the buy-in is not too difficult” [TS FG]. Rather than being invited to participate in SHIFT, HCAs on two teams reporting being told to do so. A diary entry from the initial team orientation meeting states that “when asked ‘Why are we here today?’, one CA answered, ‘I don’t know why I am here, I was just told to be here.’ Most of the CA attendees don’t show much enthusiasm about the meeting” [QA diary].

It was also evident from the focus group data that two teams did not have the protected time needed to plan and execute SHIFT PDSA cycles which diminished HCAs’ change efficacy. So, although our approach to building change efficacy during the senior sponsor R4C interview (figure 1, R4C1) by (a) discussing the importance of protected time, (b) asking if the organization is willing and able to provide such support, and (c) receiving the senior sponsor’s assurance was *delivered*, the actual provision of support to teams in the form of time for the HCAs to do the work was not *enacted* for two pilot teams. A HCA from one of these two teams stated, “we need more time. Not just 10 minutes, five minutes, rushing out and walk away…we need time to sit down and discuss this” [HCA FG]. When asked about resources and support received on the unit that was able to truly enact the management support dimension of R4C, one HCA said, “Yes. Yes. We were provided all the materials that we needed…And then we discussed. And the team sponsor, she mentioned about the project to everybody, not just us… And weekly we have a meeting to modify the changes, also what plan is working. Something like that. So yeah” [HCA FG].

### Level of Readiness for Change

All three pilot teams had strong levels of *change valence* and moderately strong levels of change efficacy. In terms of *change valence*, team sponsors were described at baseline as “believing in the value of quality improvement interventions” like SHIFT [SS interview]. Throughout the pilot, all participants commented on the value of any initiative that has the potential to improve care for residents.

*Change efficacy* of one team and their team sponsor was evident from a comment made by a senior sponsor during the initial interview: “I trust if I say to her, can we do this, can your team do this and she says ‘yes, they can’, I trust that”. Other comments reflected increases in HCA change efficacy during the pilot, although pilot data suggested that enhanced efficacy stemmed from successes the teams experienced during implementation and recognition teams received from their team sponsor and others. Asked how they felt about the SHIFT experience during the end of project focus group, a HCA stated:

> “Since you see the positive results, now we can see that we are empowered. And our confidence went up as healthcare workers…I think the confidence went up because as healthcare aids, we got to – they put us in [at report] – like I know we’re front-line workers already, but just to hear us and to get our feedback, and just kind of acknowledge us….SHIFT made us acknowledge ourselves. And like she said, you know, we look highly at ourselves” [HCA FG].

These findings highlight the difficulties of differentiating the effects of R4C activities versus other factors, on change efficacy.

R4C dimensions of *management support* and *facilitation* were less evident. End of pilot focus group data indicated that senior sponsors provided little of the support they committed to in the initial R4C interview – they tended to be “not at all involved” [HCA FG] and teams did not seem to know about the monetary stipend provided to sites to support PDSA work by front-line teams. However, the HCA exit survey data confirmed that HCAs were paid to attend LCs and other staff were paid to replace them on the unit, indicating that some of the resources promised by the senior sponsors (intended to promote change efficacy) were provided to teams.

Interestingly, the two senior sponsors who were unable to accurately predict the level of support and facilitation they themselves would provide, accurately predicted the level of support and facilitation that would be provided by the team sponsor they identified. The unit where the senior sponsor strongly endorsed the team sponsor demonstrated strong team sponsor support. Conversely, the unit where the senior sponsor was less enthusiastic about the team sponsor - but identified this individual because other possible sponsors were new to the facility or on leave - demonstrated low levels of team sponsor support. Box 1 provides illustrative longitudinal data for these two units.

**Box 1 Data Illustrative of Strong versus Weak Management Support & Facilitation R4C Dimensions**

**Strong Team Sponsor Support and Facilitation Predicted by Senior Sponsor**

- The initial senior sponsor interviewee explained why, from a readiness standpoint, she selected the manager/unit, “it’s a crew that’s been able to navigate [all of those transitions over the last year] and are, through my ongoing conversations with the manager, at a place where they’re actually working quite well together as a team…initially I had said to you [the researchers] ‘we can’t do this, we don’t have capacity for this’. But just knowing the context to this particular unit, I thought it might actually be a really good fit for them”.
- Quality advisor diary entries for this site support the senior sponsor’s assessment of the team sponsor’s readiness – she was described in the diary as enthusiastic, capable and while initially a bit directive and leading the team quite a bit, she did eventually fade back into the coaching role envisioned in the intervention.
- Field notes indicated that team members arrived at the team orientation meeting with a good understanding of the project (including topic selection), and what they were being asked to do. The team decided on 2 residents to focus on prior to the meeting and the team sponsor had explained the concept of collecting baseline information. HCAs commented a couple times about how organized the team sponsor was.
- At the end of pilot CA focus group, team members confirmed the positive choice of the team sponsor, while indicating that the senior sponsor may not have adequately enacted the role they committed to in their readiness interview: “[Respondent 2]: For our manager [team sponsor], he’s all throughout … he leads us. We get everything that we need from him. Our senior [sponsor], we haven’t met with them. [Respondent 1]: Like the senior on top of our manager, we haven’t seen them. We never see them because they’re behind closed doors”.
- This team sponsor noted that the promised financial stipend provided to the organization to support the project had not yet been given to the unit, “for the half hour that an employee is engaging on their own time when we do our weekly huddles, I’m having to pay that from my cost centre” [TS FG].

**Weak Team Sponsor Support and Facilitation predicted by Senior Sponsor**

- When asking about change valence (SHIFT’s potential to bring about improvement) during the initial senior sponsor interviewee, the senior sponsor was equivocal: “[interviewer] Do you think what you will gain will offset the required time and resource requirements? [Respondent]: We’ll see. I hope so [laughs]”.
- This senior sponsor did state they would be able to support the team (reflecting the change efficacy dimension of R4C) but they were not terribly enthusiastic about the selected team sponsor’s capabilities and readiness. When pressed, she explains that “there’s a whole bunch of factors that makes up the difference between a leader and a manager right. So, she’s a good manager, but that ability to engage staff and bring them into your ideas, is a different skill. You can make people do things, but you know, making them *want* to do things is totally different.” [initial SS interview]
- The senior sponsor noted in the interview that this was the only unit and manager that were available to participate due to manager turnover on other units.
- According to the QA diaries, the team seemed to be doing ok at the first SHIFT LC where they planned their first PDSA cycle but two weeks later the QA noted that, “the tone of the group …was quite different … Their enthusiasm and energy had changed to frustration and exhaustion …Through all of this [struggle with the change idea] the Team Sponsor sat in the background (not around the table where we were with the CA’s) and did not say a thing or offer support or a different way of looking at things” [QA diary].
- The end of pilot QA diary entry noted that “no check in or queries were made by the senior sponsor’ (who initially said she ‘hopes’ SHIFT would be valuable) and the group “exhibited a sense of defeat”.
- When asked about support the team received from the sponsors at the final focus group, the HCAs stated, “No, no, nothing; Absolutely, not once, not one word, not nothing” [CA FG].

Quantitative data from the IROC survey were received from all 14 HCAs and team sponsors from across the three pilot teams. SHIFT pilot participants’ baseline R4C perceptions showed (on a 5-point Likert-type agreement scale) descriptively higher scores on personal change valence (Mean=4.46, SD=0.66) than on organizational change valence (Mean=4.17, SD=0.44), senior management support (Mean=3.83, SD=0.56), and change efficacy (Mean=3.76, SD=0.69). With this small sample, individual and unit-level differences between the dimensions (not shown) were not statistically significant.

## DISCUSSION

Our results suggest that up-front activities requiring a few hours of a facility leader’s time, designed to gauge and also operationalize and promote readiness for change for care improvement interventions / innovations, were both feasible and acceptable and were delivered with fidelity. To promote organizational readiness for change in the pilot we utilized a ‘trickle down’ approach that focused on building readiness at the senior leadership level prior to the intervention, and then handing communication off to senior leaders, giving them strategies to build readiness among team sponsors and team members who would eventually be recruited to SHIFT. This approach seems to have helped promote readiness *at the organizational level* amongst senior leaders but revealed gaps in the enactment of the ‘trickle down’ approach.

### Lessons Learned – Modifications and Final Design

The ‘broken telephone scenario’ between project information conveyed to senior sponsors and what they, in turn, conveyed to team sponsors prompted two modifications to the *processes* (strategies) for enhancing SHIFT readiness including: (1) creating an opportunity for senior sponsors and team sponsors to attend a meeting together where they would receive the same information about SHIFT, about their respective roles, and could collaborate regarding approaches to selecting and supporting teams (this activity is shown as R4C2 in figure 1); (2) adding a second team orientation meeting (only one was included in the pilot) to enhance team member project preparation, thereby increasing change efficacy and change valence (shown as R4C3, figure 1).

Pilot results also suggested modifications to the quantitative measurement of readiness for change. Following the pilot we sought to reduce the senior sponsor R4C interview burden and strengthen R4C measurement validity. For the full implementation, we removed questions about change commitment from the initial senior sponsor interview and on the R4C questionnaire we adhered more closely to Weiner’s definition of organizational readiness for change that includes the two dimensions of change commitment and change efficacy. We added the 12-item ORIC [28] which provides a parsimonious measure of these two dimensions of R4C and we did not carry forward the personal and organizational change valence dimensions of the IROC [29] which are generally considered antecedents of readiness for change rather than dimensions of it [28,29]. All changes made between the pilot and full SHIFT intervention are summarized in Table 3.

**Table 3.** Change in R4C Processes and Assessment following the Pilot.

| **Change** | **Rationale** |
| --- | --- |
| To provide important R4C information to team and senior sponsors together, Senior Sponsors were invited to attend a session together with their Team Sponsor (figure 1, R4C2/LS1) | To promote role clarity and improve enactment of certain R4C elements – addresses the gap between R4C information delivered to senior sponsors and R4C information they conveyed to team sponsors. |
| Instead of one 30-minute onsite team orientation meeting prior to the first SHIFT LC; the final R4C design would include two onsite meetings with teams prior to the first LC (figure 1, R4C3). | To enhance change efficacy and change valence by providing more time for (a) project orientation and (b) discussion of the QI topic teams would work on. |
| Senior Sponsor R4C interview (figure 1, R4C1) was reduced from 90 minutes to 60 minutes; questions regarding change commitment were removed from the interview and added to the R4C survey. | To reduce senior sponsor interview burden. |
| IROC change valence dimensions were removed; the 12-item ORIC which measures core R4C dimensions (change commitment and change efficacy) were added | To reduce survey burden at SHIFT LCs and ensure the R4C measures is parsimonious and provided a valid reflection of the key construct dimensions [8,28] |
| R4C was measured longitudinally, at the start of SHIFT, the mid-point, and at the final LC | To be able to explore change efficacy enhancement processes |

### Theoretical and Measurement Implications

We concur with Weiner et al., [8] that while readiness for change is a multi-level construct, it is best conceived beyond the individual level as it reflects a shared resolve and capacity to implement process changes that typically involve a complex network of workers, teams and leaders and so reflects an organization’s collective willingness (change commitment) and ability (change efficacy) to implement a change. To promote greater information exchange and role clarity, thereby directly building change commitment and change efficacy (the core dimensions of R4C), we suggest carrying out readiness for change activities with all of those involved *across an organization* who have a role to play in the change initiative.

Although we found that the team with stronger baseline readiness seemed to be more successful at building readiness among the team sponsor and team members, our results suggest it is challenging to parse out the consequences of readiness for change activities on change implementation. Our pilot R4C activities were intended to promote change valence, build management support, and promote facilitation and, in turn, to enhance change commitment and change efficacy. However, management support and facilitation are also core components of the SHIFT intervention and are therefore key indicators of intervention implementation / enactment. So, if the anticipated management support does not materialize, it is not clear if this is a R4C enactment problem or an intervention enactment problem. This conundrum underscores the importance of adhering to Weiner’s definition of R4C that includes only the dimensions of change commitment and efficacy - with change valence as an antecedent of change commitment. Management support and facilitation would, using a similar argument, also best be considered as antecedents of change efficacy. Indeed, Weiner [8] and Shea [28] argue that efficacy reflects a psychological state but that it is a function of people’s perceptions of (a) more structural things including whether they have the resources necessary to implement a change, and (b) whether situational factors such as timing and, presumably, support are seen as favourable.

Relatedly, while enhanced change efficacy is a core element of readiness for change, our pilot results, and the results of prior work in SCOPE [5,6,20], clearly indicate that change efficacy is relevant beyond the initial ‘readiness for change’ period. Indeed, we found change efficacy *developed* as teams implemented the intervention and, in particular, when they witnessed the intervention having positive effects on residents. These findings are consistent with social cognitive theory indicating that efficacy beliefs are formed from feedback received during the change process such that efficacy judgments increase when a change appears to have the intended positive effects, and may decrease when a change does not appear to be working (depending on attributions made) [31,32].

This raises non-trivial theoretical questions pertaining to the roots of change efficacy. Drawing on foundational work by Gist and Mitchell (1992) [32], the theory of organizational readiness for change [8] emphasizes material supports that organizational members feel they have, and their assessment of task demands and situational factors as factors that influence change efficacy. However, foundational work in social cognitive theory on change efficacy [33] (including Gist and Mitchell [3 2]) suggests feedback regarding the success or failure of the change process (referred to as personal experience with mastery) is a highly influential source of efficacy judgements. The Theory of organizational readiness for change may therefore benefit from a broader, more longitudinal conceptualization that recognizes efficacy judgements “change over time as new information and experience are acquired” [32], and that feedback on performance attainment plays a central role in influencing collective efficacy during the change process [34]. This is an area worthy of further investigation as are questions which have been raised regarding whether the formation of efficacy judgements is isomorphic at the individual and group levels [34]. R4C is a multi-level construct so it is prudent to explore whether the same processes influence self-efficacy judgements and collective efficacy judgements. In the full SHIFT intervention, we opted to measure R4C longitudinally to be able to more completely explore these efficacy enhancement questions. As the field refines R4C theory, such questions regarding the roots of efficacy judgements are important to investigate given the robust body of evidence regarding social cognitive theory which shows efficacy beliefs contribute significantly to motivation and performance.

In terms of assessing readiness for change, our study supports the value of exploring this complex construct using both qualitative and quantitative approaches to provide a nuanced understanding of how readiness for change manifests longitudinally as a change initiative unfolds. Our work to operationalize and evaluate readiness for change contributes to a substantial body of existing work on the measurement of R4C [35,36] and its relationship to a range of outcomes [10,11,37]. The more substantive contribution may be to the small but growing body of literature on how to promote/build readiness for change [12,13,38].

## CONCLUSION

Implementation Science is focused on making change initiatives work. R4C is recognized as an important implementation determinant and the current paper contributes to a limited knowledge base regarding how to operationalize this construct to promote health system change and improvement. Informed by this pilot work, a larger scale roll out of the R4C module in 10 sites is underway as part of the full-scale SHIFT type III hybrid implementation-effectiveness trial. The SHIFT trial will shed light on (i) the mechanisms by which readiness for change (including its components of change efficacy and change commitment) are strengthened and influence implementation, and (ii) how to build and sustain partnerships between researchers, care delivery sites and policy decision makers in a way that primes the system and organizations and promotes readiness for change.

## Data Availability

Data generated and/or analysed during the current study are not publicly available in accordance with the privacy and confidentiality requirements of the University of Manitoba Research Ethics Board. Select data may be available from the corresponding author on reasonable request.

## LIST OF ABBREVIATIONS

FAP: Facilitated Action Period
IHI: Institute for Healthcare Improvement
TM: Team member
TS: Team sponsor (unit manager)
NH: Nursing Home
PDSA: Plan-Do-Study-Act cycle
QA: Quality Advisor
QI: Quality Improvement
R4C: Readiness for Change
SCOPE: ‘**S**afer **C**are for **O**lder **P**ersons (in residential) **E**nvironments’ (Intervention)
SHIFT: **S**upporting **H**ealthcare **I**mprovement Through **F**acilitation and **T**raining (Intervention)
SS: Senior Sponsor (facility senior leader)

## DECLARATIONS

### Ethics approval and consent to participate

This study was reviewed and approved by the University of Manitoba (Bannatyne Campus) Health Research Ethics Board (HREB), reference number: HS18486 (H2015:045). Written consent was obtained from all study participants.

### Consent for publication

N/A

### Competing interests

The authors declare that they have no competing interests

### Funding

SHIFT is part of the larger Supporting Older Adult Healthcare Reform through Research (SOARR) research program, funded by Manitoba Health, Seniors and Long-Term Care (MHSLTC) through a Service Purchase Agreement between the Government of Manitoba and the University of Manitoba. The results of this study are those of the authors, and no official endorsement by MHSLTC is intended or should be inferred.

### Authors’ contributions

MBD, LK and LG lead the development of the R4C module. LR and GV contributed to design feasibility. MBD, LG, WB, MH, CE and AW conceived and refined SHIFT design and measurement. LK, JP, and DS operationalized all day-to-day aspects of LeaderSHIFT, including data collection. LG prepared the initial draft of this manuscript, and all authors reviewed and approved the final manuscript version.

## Acknowledgements

We wish to thank Don McLeod (SHIFT Senior Quality Advisor) and Cristie Perfas (SHIFT Quality Advisor) for their work and dedication to the QI teams and their team sponsors. We are also grateful to those who participated in and enabled this study – thank you for your engagement and feedback.

## Notes

### Competing Interest Statement

The authors have declared no competing interest.

### Clinical Trial

Trial ID: NCT03426072

### Author Declarations

This study was reviewed and approved by Health Research Ethics Board (HREB) of the University of Manitoba (Bannatyne Campus) who gave ethical approval for this work (Ref number: HS18486 (H2015:045)).

